# Association between afterload-related cardiac performance and acute kidney injury in patients with sepsis: a retrospective cohort study

**DOI:** 10.1101/2024.12.10.24318824

**Authors:** Jie-zhao Zheng, Ze-bin Guo, Li Li, Hong-xuan Zhou, Jun-yu Huang, Min Luo, Min-xuan Huang, Jia-xian Huo, Wen-xing Huang, Hao-ran Yuan, Hong-xi Chen, Zi-xin Jiang, Wei-yan Chen

## Abstract

**Background:** It is believed that there is a complex bidirectional causal and pathophysiological relationship between heart and kidney in sepsis. Afterload-related cardiac performance (ACP) has been found to have a close association with cardiac function and prognosis in sepsis. The aim of this study was to explore its correlation with sepsis-associated acute kidney injury (SA-AKI).

**Methods:** 150 patients with sepsis who underwent PiCCO were included. Restricted cubic spline (RCS) and Cox proportional hazards regression were used to describe the relationship between ACP and SA-AKI. The receiver operating characteristic curve (AUROC) analysis was used to evaluate the predictive value of ACP for SA-AKI. A prediction model of SA-AKI occurrence probability within 7 days was presented by nomogram.

**Results:** 67.3% patients developed SA-AKI within 7 days. The RCS model demonstrated that the risk of SA-AKI increased with the decrease of ACP. Cox regression analyses also found an independent inverse association between them (HR 0.974, 95%CI 0.963 - 0.985, p < 0.001). ACP showed significantly greater discrimination for SA-AKI than CI (*p* = 0.004). Independent risk factors for SA-AKI within 7 days were history of hypertension, type of pathogens, SOFA score, serum potassium and ACP. The AUROC of the nomogram established by age and the above factors was 0.865 (95%CI 0.808 – 0.923, *p* < 0.001) and the C-index was 0.800 (95%CI 0.753 - 0.847).

**Conclusions:** Low ACP was associated with an increased risk of SA-AKI within 7 days. ACP, which might be superior to CI, had the potential for early prediction of SA-AKI.

## Introduction

Acute kidney injury (AKI) is a common clinical syndrome characterized by a rapid decline in renal function within a short period of time^[1]^. Sepsis is the major cause of AKI in critical ill patients, accounting for 25% to 75% of all cases^[2–4]^. According to studies, sepsis-associated AKI (SA-AKI) is strongly associated with worse outcomes, including longer intensive care unit (ICU) and hospital stays, higher mortality, an increased risk of chronic kidney diseases (CKD) and reduced quality of life^[5–8]^. Therefore, early identification and intervention of SA-AKI are of great significance for improving the prognosis of patients.

Creatinine, as a classic indicator of renal function, has a long half-life and is influenced by a variety of factors, including muscle mass, physical activity, dietary habits and age, which can lead to delayed recognition of renal dysfunction^[1, 9]^. It is reported that numerous novel biomarkers such as neutrophil gelatinase associated lipocalcin (NGAL) predict SA-AKI with high accuracy. However, the application of these forecasting biomarkers is still limited by the promotion of tests, costs and further validation of their clinical significance is needed^[10–15]^.

The pathogenesis of SA-AKI is complex. It is hypothesized that SA-AKI may be the result of a combination of mechanisms such as inflammatory injury, mitochondrial dysfunction, abnormal renal perfusion, microcirculatory dysfunction and abnormalities^[1, 5, 16]^. Besides, recent studies have shown that early renal function changes in sepsis (within the first 48 hours) appear to represent primarily a functional rather than a structural disease^[17–19]^. Avoidance of nephrotoxins, optimization of fluid and hemodynamic status are currently limited means to prevent the occurrence or progression of SA-AKI and to protect renal function^[1]^. This, in turn, suggests that the abnormality of renal perfusion may be the initiating factor of AKI, and early hemodynamic indicators may potentially signal the onset and progression of AKI at an earlier stage^[18]^.

The heart is also one of the most commonly involved organs in sepsis. Sepsis-induced myocardial dysfunction (SIMD) can lead to more severe hemodynamic disorder, which subsequently leads to abnormal renal perfusion pressure (RPP) and an increase in the incidence of SA-AKI. Clinical studies have shown that some hemodynamic indicators, such as mean arterial pressure (MAP) and cardiac index (CI), are risk factors affecting the mortality of patients in ICU with AKI. While adequate RRP, MAP and cardiac output (CO) were related to the prevention of SA-AKI^[20–22]^. Therefore, there is a theoretical possibility to identify SA-AKI as early as possible through hemodynamic indicators.

Afterload-related cardiac performance (ACP), first introduced by Werdan et al. in 2011, is a ratio of measured CO to predicted CO, which presents the cardiac ability to increase its output when systemic vascular resistance (SVR) decreases in order to maintain a constant MAP^[23]^. Since it takes into account the influence of MAP, SVR on CO, it may be more advantageous in quantitatively measuring of SIMD and predicting SA-AKI.

Our study aims to investigate the relationship between ACP and SA-AKI, and further verify its predictive value, so as to provide a clinical method for the prediction of SA-AKI.

## Methods

### Study population and setting

This study was a retrospective cohort study involving adult Patients (aged ≥ 18 years) who were diagnosed with sepsis and underwent PiCCO during their ICU stay between June 2016 and December 2021. Sepsis was defined as organ dysfunction, identified by an acute change of ≥ 2 points in the total Sequential Organ Failure Assessment (SOFA) score, caused by the suspected or confirmed infection (Sepsis-3)^[24]^. Fluid resuscitation and antibiotic therapy had been administered according to guideline^[25]^. Moreover, SA-AKI was diagnosed by meeting the Kidney Disease: Improving Global Outcomes (KDIGO) criteria for AKI^[26]^, in the presence of sepsis^[16]^. Patients were excluded if they met any of the following criteria: (1) repeat ICU admissions from the same hospital episode; (2) the length of ICU stay was less than 24 hours; (3) previous history of congenital heart disease, severe valvular heart disease; (4) history of CKD, or renal replacement has been performed; (5) AKI had occurred before or at the time of admission to the ICU; (6) main diagnoses directly related to myocardial dysfunction, such as acute myocardial infarction, myocarditis, or moderate-to-severe pericardial effusion; (7) post-cardiopulmonary resuscitation status.

### Ethical approval

The Clinical Research and Application Institutional Review Board of the Second Affiliated Hospital of Guangzhou Medical University have approved this study (number: 2023-hg-ks-07). Since the nature of the retrospective observational study and the data were anonymized at the time of collection which did not involve privacy, The need for individual informed consent was waived.

### Afterload-related cardiac performance (ACP)

ACP (%) was calculated using the formula previously described by Werdan et al.^[23]^ and was described as CO_measured_ / CO_predicted as normal_ ×100. CO_predicted as normal_ in the formula was calculated as [560.68× (MAP - CVP) × 80 / CO_measure_] ^-0.645^. CVP was an acronym for central venous pressure. ACP was classified as normal (>80%), slight impairment (60-80%), moderate-severe impairment (≤60%) to distinguish cardiac function.

### Data collection

We extracted the following variables: demographic characteristics, vital signs, laboratory tests, interventions [using of mechanical ventilation, continuous renal replacement therapy (CRRT) during ICU stay] and history of comorbidities [chronic heart failure (CHF), atrial fibrillation (AF), hypertension and diabetics]. Primary diagnosis and pathogenic culture results were also collected. The severity of the diseases was estimated using SOFA scores, and acute physiology and chronic health evaluation (APACHE) II score. If vasoactive drugs were used, the vasoactive inotropic score (VIS) was calculated based on the maximum dose on the first day of sepsis^[27]^. Parameters, such as CO, CI, cardiac power index (CPI), global end-diastolic volume index (GEDI), systemic vascular resistance index (SVRI), extravascular lung water index (ELWI), CVP and MAP, were recorded after PiCCO monitoring. RPP was calculated as (MAP – CVP)^[28]^.

Variables missing more than 20% were removed from this analysis, and the remaining variables were imputed via the method of multiple imputation.

### Outcomes

The primary outcome was to investigate the association between ACP and risk of SA-AKI within 7 days, and further explore the value of ACP in predicting the 7-day SA-AKI occurrence probability.

### Statistical analysis

All patients were divided into three groups according to ACP classification of cardiac function. A descriptive analysis was performed. The chi-square test, One-Way ANOVA, Kruskal-Wallis test were conducted for group comparisons of categorical, normally distributed, and nonnormally distributed continuous variables, respectively.

A Model fitted with restricted cubic spline was conducted to test for linear or nonlinear shapes of the association between ACP and the probability of SA-AKI. Collinearity analysis was performed before multivariate Cox regression analysis, and variables with variance inflation factor (VIF) > 5 were excluded. Cox proportional hazards regression analyses were used to evaluate the independent associations between ACP and SA-AKI after adjusting for confounding factors, and further establish a nomogram prediction model. Internal validation was performed using 500 bootstrap and calibration curves were drawn. The concordance index (C-index), area under the curve (AUC) and 95% confidence interval (95%CI) were calculated. The receiver operating characteristic curve (ROC) was used to assess the efficacy of ACP in predicting the risk of SA-AKI, and then the predictive ability of ACP with CI, CPI and RPP for SA-AKI was tested by comparing the AUC. For survival analysis, Kaplan-Meier curves were depicted and compared by log-rank test.

A two-tailed test was performed and *p* < 0.05 was considered statistically significant. All analyses were performed using the statistical software packages R. version 4.4.1 (R Foundation for Statistical Computing, Vienna, Austria) and Free Statistics software versions 1.9.

## Results

### Baseline characteristics

Between June 2016 and December 2021, a total of 1486 individuals with sepsis according to the Sepsis-3.0 criterion, of which 637 patients were underwent PiCCO in ICU, and 150 patients were finally enrolled in this study. The flow chart of the study participants selection was presented in Figure 1.

**Figure 1.**
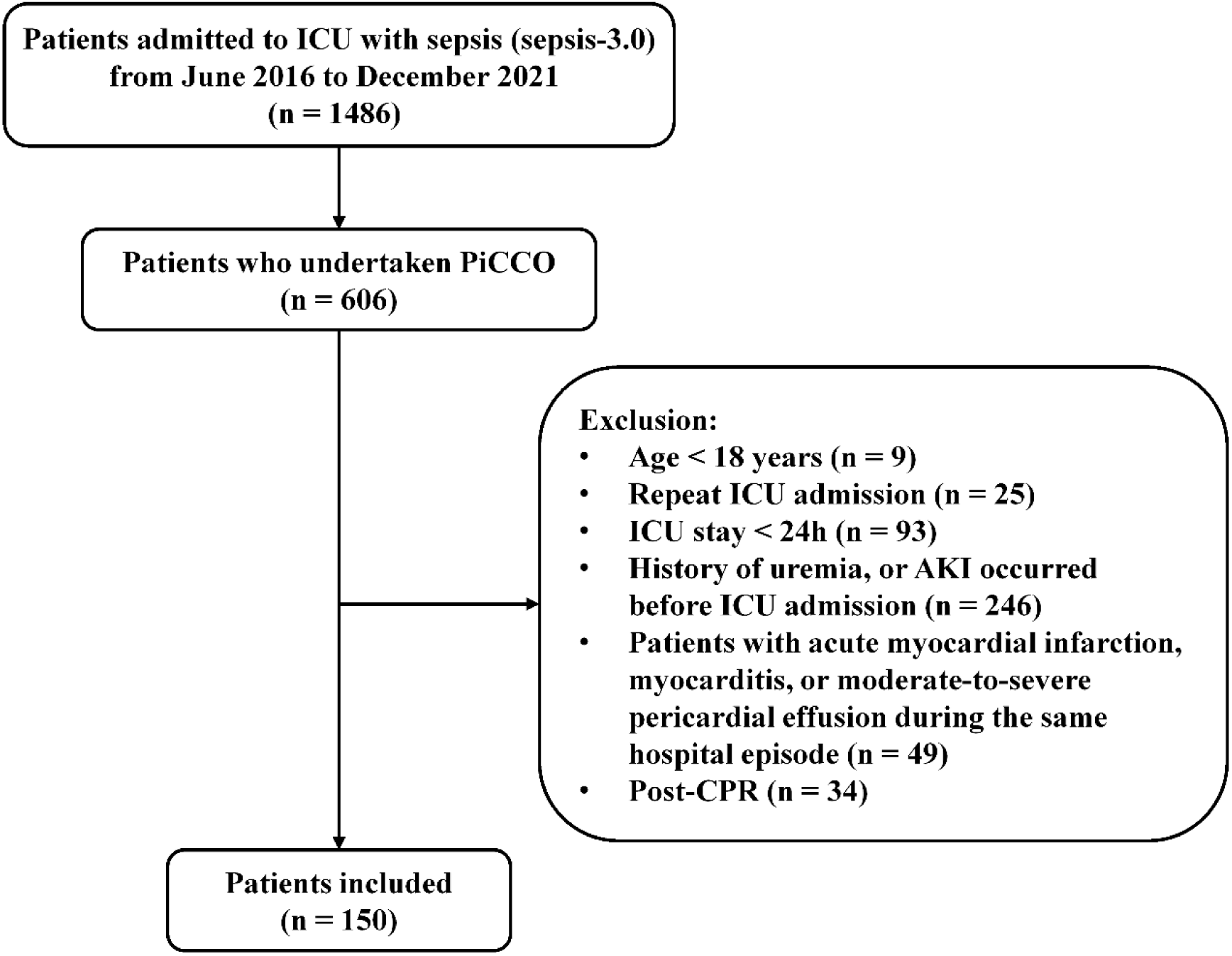
The flow chart of the study.

In this cohort, the predominant site of infection was the lungs, followed by the abdomen. Among these cases, 71.3% patients progressed to septic shock, with nearly all patients required ventilator support, and 67.3% patients developing AKI within 7 days following diagnosis of sepsis. Of the patients who developed SA-AKI, 68.3% patients received CRRT. patients with AKI had lower ACP compared to those without AKI (*p* < 0.001) (Table 1). Upon grouping patients by ACP level, it was observed that patients with moderate-severe myocardial impairment were characterized by older, higher SOFA scores and VIS, higher level of CVP and serum lactate, lower MAP, and greater need for CRRT. Moreover, the incidence of AKI increased with the severity of cardiac functional impairment, and there was statistical difference between groups (*p* < 0.001, Table S1).

**Table 1.**
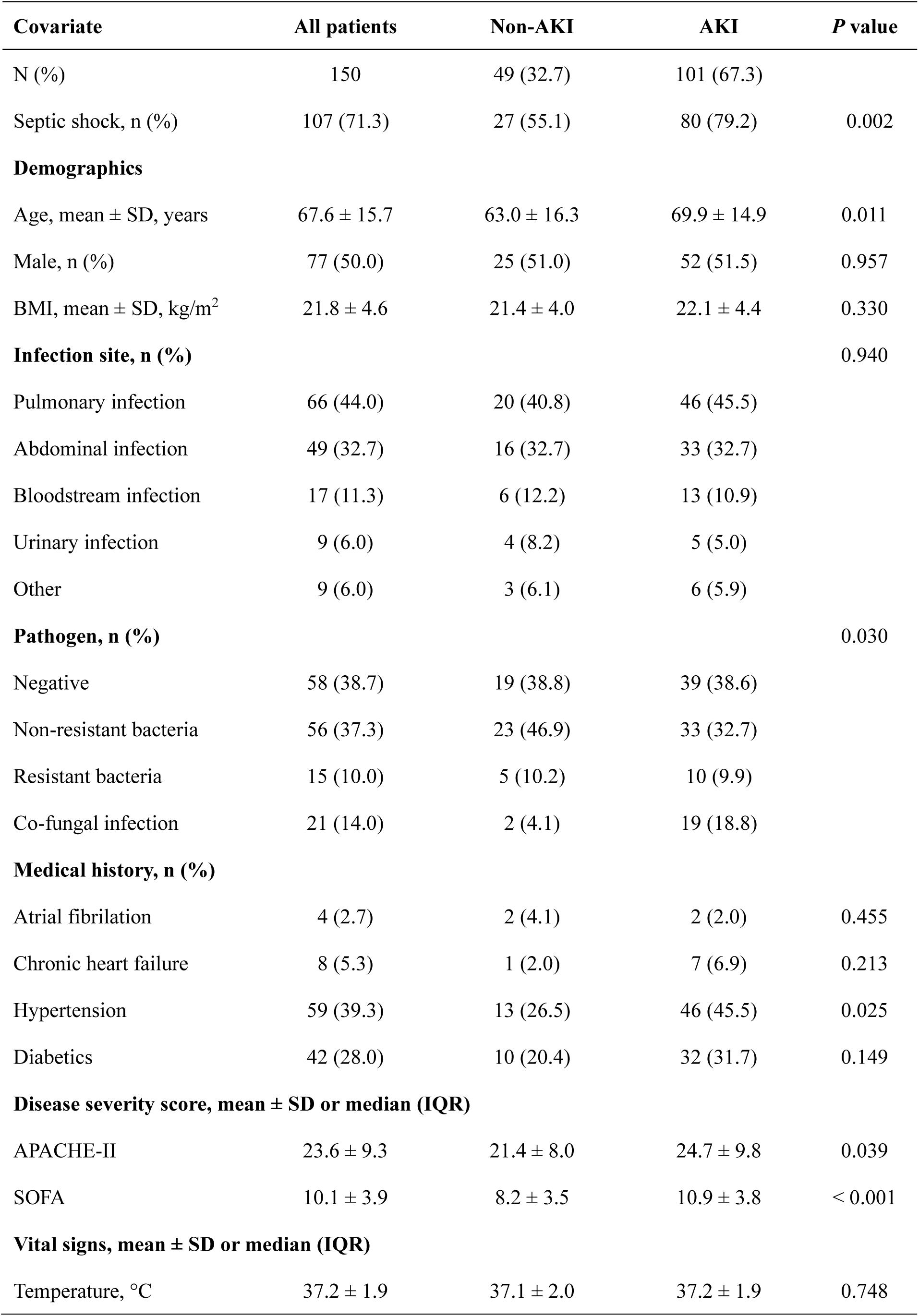
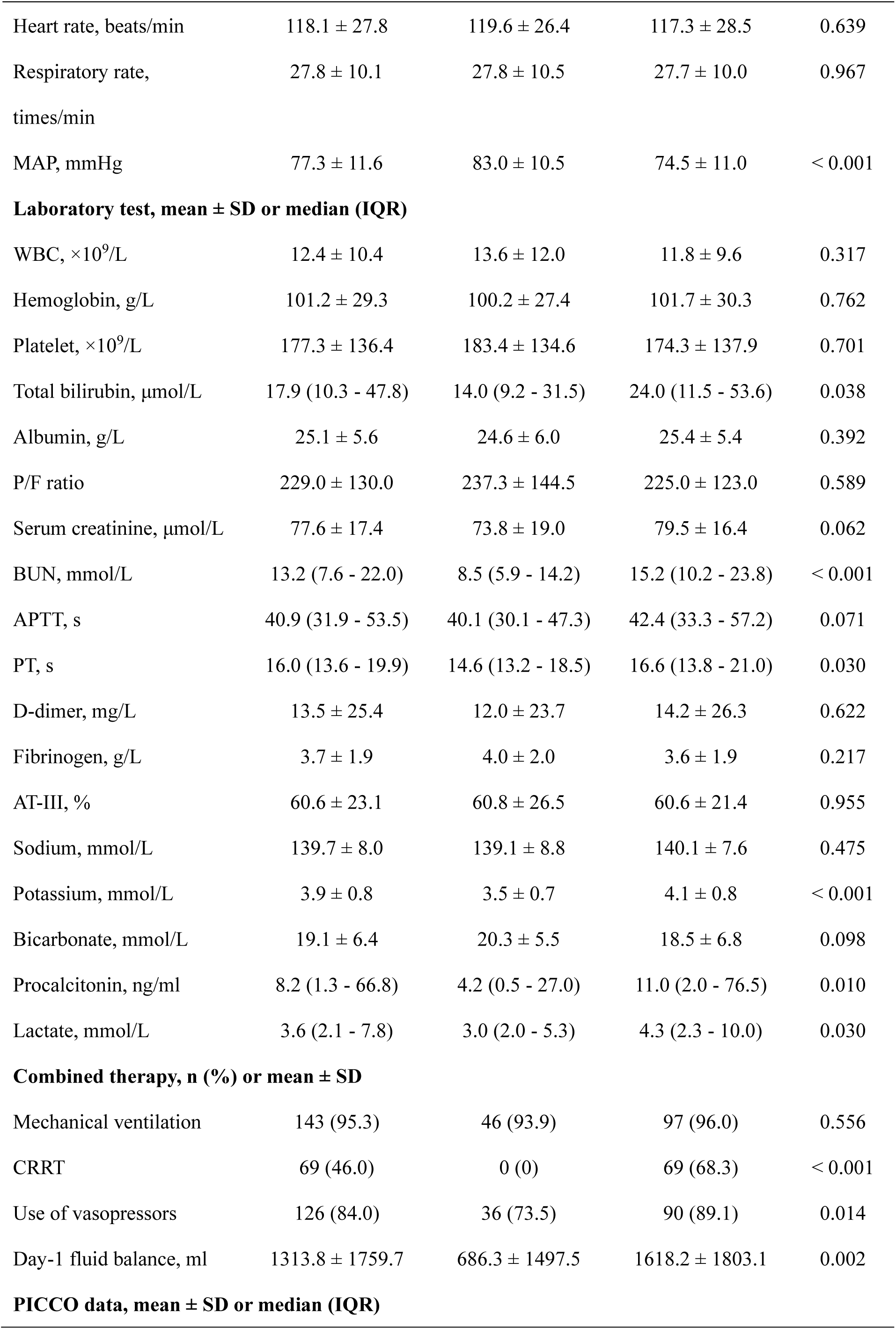
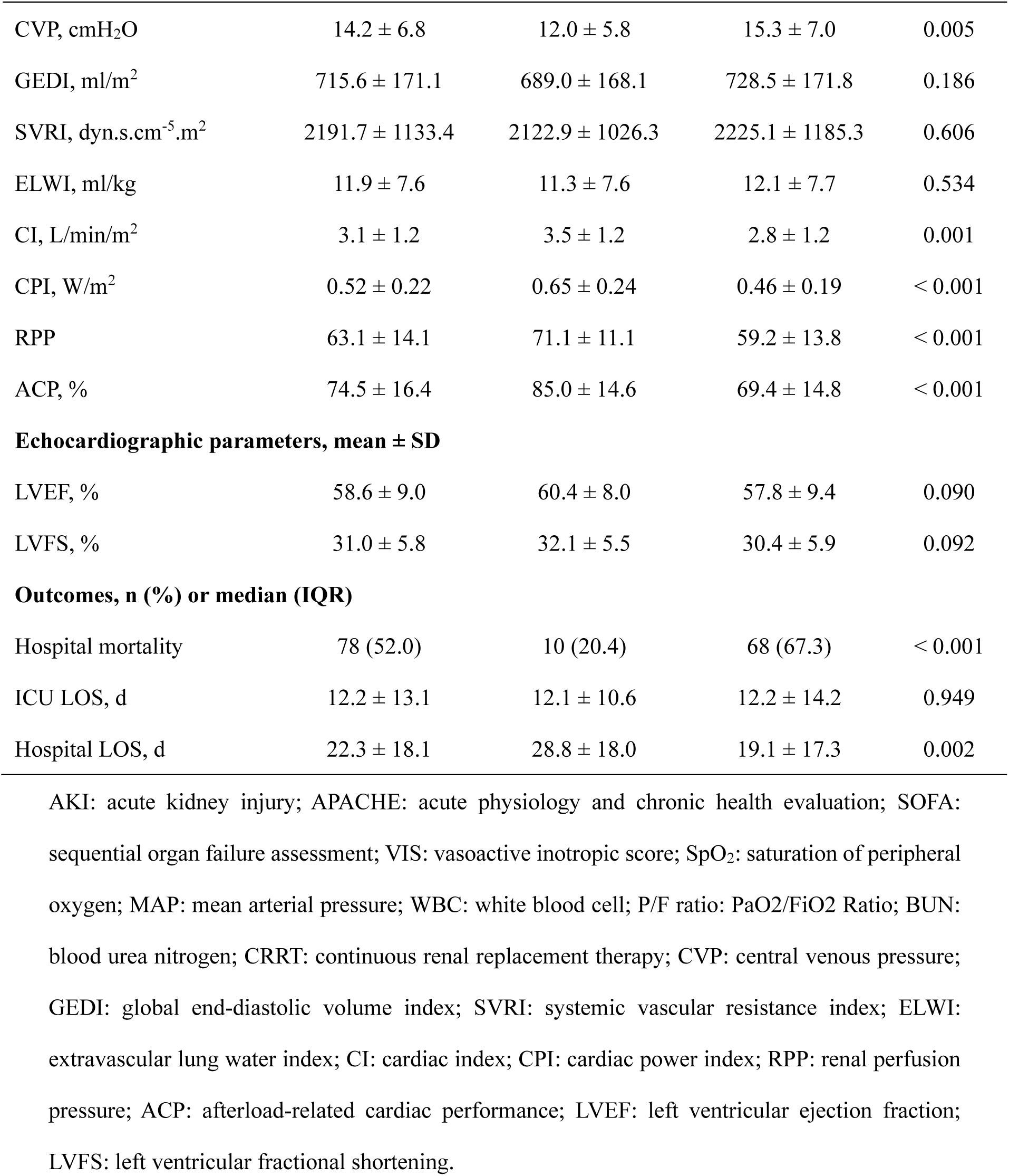
Baseline characteristics of AKI and non-AKI patients.

| **Covariate** | **All patients** | **Non-AKI** | **AKI** | ***P* value** |
| --- | --- | --- | --- | --- |
| N (%) | 150 | 49 (32.7) | 101 (67.3) |  |
| Septic shock, n (%) | 107 (71.3) | 27 (55.1) | 80 (79.2) | 0.002 |
| **Demographics** |  |  |  |  |
| Age, mean ± SD, years | 67.6 ± 15.7 | 63.0 ± 16.3 | 69.9 ± 14.9 | 0.011 |
| Male, n (%) | 77 (50.0) | 25 (51.0) | 52 (51.5) | 0.957 |
| BMI, mean ± SD, kg/m<sup>2</sup> | 21.8 ± 4.6 | 21.4 ± 4.0 | 22.1 ± 4.4 | 0.330 |
| **Infection site, n (%)** |  |  |  | 0.940 |
| Pulmonary infection | 66 (44.0) | 20 (40.8) | 46 (45.5) |  |
| Abdominal infection | 49 (32.7) | 16 (32.7) | 33 (32.7) |  |
| Bloodstream infection | 17 (11.3) | 6 (12.2) | 13 (10.9) |  |
| Urinary infection | 9 (6.0) | 4 (8.2) | 5 (5.0) |  |
| Other | 9 (6.0) | 3 (6.1) | 6 (5.9) |  |
| **Pathogen, n (%)** |  |  |  | 0.030 |
| Negative | 58 (38.7) | 19 (38.8) | 39 (38.6) |  |
| Non-resistant bacteria | 56 (37.3) | 23 (46.9) | 33 (32.7) |  |
| Resistant bacteria | 15 (10.0) | 5 (10.2) | 10 (9.9) |  |
| Co-fungal infection | 21 (14.0) | 2 (4.1) | 19 (18.8) |  |
| **Medical history, n (%)** |  |  |  |  |
| Atrial fibrillation | 4 (2.7) | 2 (4.1) | 2 (2.0) | 0.455 |
| Chronic heart failure | 8 (5.3) | 1 (2.0) | 7 (6.9) | 0.213 |
| Hypertension | 59 (39.3) | 13 (26.5) | 46 (45.5) | 0.025 |
| Diabetics | 42 (28.0) | 10 (20.4) | 32 (31.7) | 0.149 |
| **Disease severity score, mean ± SD or median (IQR)** |  |  |  |  |
| APACHE-II | 23.6 ± 9.3 | 21.4 ± 8.0 | 24.7 ± 9.8 | 0.039 |
| SOFA | 10.1 ± 3.9 | 8.2 ± 3.5 | 10.9 ± 3.8 | < 0.001 |
| **Vital signs, mean ± SD or median (IQR)** |  |  |  |  |
| Temperature, °C | 37.2 ± 1.9 | 37.1 ± 2.0 | 37.2 ± 1.9 | 0.748 |
| Heart rate, beats/min | 118.1 ± 27.8 | 119.6 ± 26.4 | 117.3 ± 28.5 | 0.639 |
| Respiratory rate,<br>times/min | 27.8 ± 10.1 | 27.8 ± 10.5 | 27.7 ± 10.0 | 0.967 |
| MAP, mmHg | 77.3 ± 11.6 | 83.0 ± 10.5 | 74.5 ± 11.0 | < 0.001 |
| **Laboratory test, mean ± SD or median (IQR)** |  |  |  |  |
| WBC, ×10<sup>9</sup>/L | 12.4 ± 10.4 | 13.6 ± 12.0 | 11.8 ± 9.6 | 0.317 |
| Hemoglobin, g/L | 101.2 ± 29.3 | 100.2 ± 27.4 | 101.7 ± 30.3 | 0.762 |
| Platelet, ×10<sup>9</sup>/L | 177.3 ± 136.4 | 183.4 ± 134.6 | 174.3 ± 137.9 | 0.701 |
| Total bilirubin, μmol/L | 17.9 (10.3 - 47.8) | 14.0 (9.2 - 31.5) | 24.0 (11.5 - 53.6) | 0.038 |
| Albumin, g/L | 25.1 ± 5.6 | 24.6 ± 6.0 | 25.4 ± 5.4 | 0.392 |
| P/F ratio | 229.0 ± 130.0 | 237.3 ± 144.5 | 225.0 ± 123.0 | 0.589 |
| Serum creatinine, μmol/L | 77.6 ± 17.4 | 73.8 ± 19.0 | 79.5 ± 16.4 | 0.062 |
| BUN, mmol/L | 13.2 (7.6 - 22.0) | 8.5 (5.9 - 14.2) | 15.2 (10.2 - 23.8) | < 0.001 |
| APTT, s | 40.9 (31.9 - 53.5) | 40.1 (30.1 - 47.3) | 42.4 (33.3 - 57.2) | 0.071 |
| PT, s | 16.0 (13.6 - 19.9) | 14.6 (13.2 - 18.5) | 16.6 (13.8 - 21.0) | 0.030 |
| D-dimer, mg/L | 13.5 ± 25.4 | 12.0 ± 23.7 | 14.2 ± 26.3 | 0.622 |
| Fibrinogen, g/L | 3.7 ± 1.9 | 4.0 ± 2.0 | 3.6 ± 1.9 | 0.217 |
| AT-III, % | 60.6 ± 23.1 | 60.8 ± 26.5 | 60.6 ± 21.4 | 0.955 |
| Sodium, mmol/L | 139.7 ± 8.0 | 139.1 ± 8.8 | 140.1 ± 7.6 | 0.475 |
| Potassium, mmol/L | 3.9 ± 0.8 | 3.5 ± 0.7 | 4.1 ± 0.8 | < 0.001 |
| Bicarbonate, mmol/L | 19.1 ± 6.4 | 20.3 ± 5.5 | 18.5 ± 6.8 | 0.098 |
| Procalcitonin, ng/ml | 8.2 (1.3 - 66.8) | 4.2 (0.5 - 27.0) | 11.0 (2.0 - 76.5) | 0.010 |
| Lactate, mmol/L | 3.6 (2.1 - 7.8) | 3.0 (2.0 - 5.3) | 4.3 (2.3 - 10.0) | 0.030 |
| **Combined therapy, n (%) or mean ± SD** |  |  |  |  |
| Mechanical ventilation | 143 (95.3) | 46 (93.9) | 97 (96.0) | 0.556 |
| CRRT | 69 (46.0) | 0 (0) | 69 (68.3) | < 0.001 |
| Use of vasopressors | 126 (84.0) | 36 (73.5) | 90 (89.1) | 0.014 |
| Day-1 fluid balance, ml | 1313.8 ± 1759.7 | 686.3 ± 1497.5 | 1618.2 ± 1803.1 | 0.002 |
| **PICCO data, mean ± SD or median (IQR)** |  |  |  |  |
| CVP, cmH<sub>2</sub>O | 14.2 ± 6.8 | 12.0 ± 5.8 | 15.3 ± 7.0 | 0.005 |
| GEDI, ml/m<sup>2</sup> | 715.6 ± 171.1 | 689.0 ± 168.1 | 728.5 ± 171.8 | 0.186 |
| SVRI, dyn.s.cm<sup>-5</sup>.m<sup>2</sup> | 2191.7 ± 1133.4 | 2122.9 ± 1026.3 | 2225.1 ± 1185.3 | 0.606 |
| ELWI, ml/kg | 11.9 ± 7.6 | 11.3 ± 7.6 | 12.1 ± 7.7 | 0.534 |
| CI, L/min/m<sup>2</sup> | 3.1 ± 1.2 | 3.5 ± 1.2 | 2.8 ± 1.2 | 0.001 |
| CPI, W/m<sup>2</sup> | 0.52 ± 0.22 | 0.65 ± 0.24 | 0.46 ± 0.19 | < 0.001 |
| RPP | 63.1 ± 14.1 | 71.1 ± 11.1 | 59.2 ± 13.8 | < 0.001 |
| ACP, % | 74.5 ± 16.4 | 85.0 ± 14.6 | 69.4 ± 14.8 | < 0.001 |
| **Echocardiographic parameters, mean ± SD** |  |  |  |  |
| LVEF, % | 58.6 ± 9.0 | 60.4 ± 8.0 | 57.8 ± 9.4 | 0.090 |
| LVFS, % | 31.0 ± 5.8 | 32.1 ± 5.5 | 30.4 ± 5.9 | 0.092 |
| **Outcomes, n (%) or median (IQR)** |  |  |  |  |
| Hospital mortality | 78 (52.0) | 10 (20.4) | 68 (67.3) | < 0.001 |
| ICU LOS, d | 12.2 ± 13.1 | 12.1 ± 10.6 | 12.2 ± 14.2 | 0.949 |
| Hospital LOS, d | 22.3 ± 18.1 | 28.8 ± 18.0 | 19.1 ± 17.3 | 0.002 |

AKI: acute kidney injury; APACHE: acute physiology and chronic health evaluation; SOFA: sequential organ failure assessment; VIS: vasoactive inotropic score; SpO<sub>2</sub>: saturation of peripheral oxygen; MAP: mean arterial pressure; WBC: white blood cell; P/F ratio: PaO<sub>2</sub>/FiO<sub>2</sub> Ratio; BUN: blood urea nitrogen; CRRT: continuous renal replacement therapy; CVP: central venous pressure; GEDI: global end-diastolic volume index; SVRI: systemic vascular resistance index; ELWI: extravascular lung water index; CI: cardiac index; CPI: cardiac power index; RPP: renal perfusion pressure; ACP: afterload-related cardiac performance; LVEF: left ventricular ejection fraction; LVFS: left ventricular fractional shortening.

### Predictive value of ACP, CI, CPI and RPP

Our findings demonstrated that ACP, CI, CPI and RPP exhibited a high rate of identification of SA-AKI, with statistically significant differences (*p* < 0.001) (Table 2). The AUC value of ACP at enrollment was the highest. ACP at enrollment significantly outperformed CI in predicting SA-AKI within 7 days (*p* = 0.004) (Table S2). The AUC value of ACP, while not demonstrating a statistically significant improvement over that of CPI and RPP in predicting the occurrence of SA-AKI within 7 days, achieved the highest value at 0.777.

**Table 2.** Discriminative abilities of different indicators to assess SA-AKI.

| **Variables** | **AUC** | **95%CI** | **Cut-off**<br>**value** | **Sensitivity**<br>**(%)** | **Specificity**<br>**(%)** | **Accuracy**<br>**(%)** | ***P***<br>**value** |
| --- | --- | --- | --- | --- | --- | --- | --- |
| **ACP-0h (%)** | 0.777 | 0.696 - 0.857 | 78.6 | 74.3 | 71.4 | 73.3 | <0.001 |
| **ACP-6h (%)** | 0.774 | 0.694 - 0.853 | 74.3 | 64.2 | 83.7 | 70.8 | <0.001 |
| **ACP-12h (%)** | 0.725 | 0.638 - 0.811 | 77.0 | 58.1 | 87.2 | 68.4 | <0.001 |
| **ACP-18h (%)** | 0.673 | 0.582 - 0.764 | 73.2 | 51.1 | 78.7 | 60.4 | <0.001 |
| **ACP-24h (%)** | 0.706 | 0.617 - 0.794 | 77.7 | 62.4 | 78.7 | 68.2 | <0.001 |
| **CPI-0h (W/m<sup>2</sup>)** | 0.738 | 0.650 - 0.826 | 0.58 | 76.2 | 65.3 | 72.7 | <0.001 |
| **CI-0h (L/min/m<sup>2</sup>)** | 0.678 | 0.585 - 0.770 | 3.1 | 67.3 | 67.3 | 67.3 | <0.001 |
| **RPP-0h (mmHg)** | 0.742 | 0.662 - 0.822 | 60.4 | 55.4 | 87.8 | 66.0 | <0.001 |

ACP: Afterload-related cardiac performance; RPP: Renal perfusion pressure; CPI: Cardiac power index; CI: Cardiac index;

### Correlation between ACP and SA-AKI

To further investigate the association between ACP and SA-AKI, we employed restricted cubic splines flexible model to visualize the relationship between ACP and SA-AKI risk (Figure 2). The findings indicated that ACP < 75.8% was identified as a risk factor for SA-AKI, and the risk of developing SA-AKI escalated rapidly with decreasing ACP levels, indicating a negative correlation between them.

**Figure 2.**
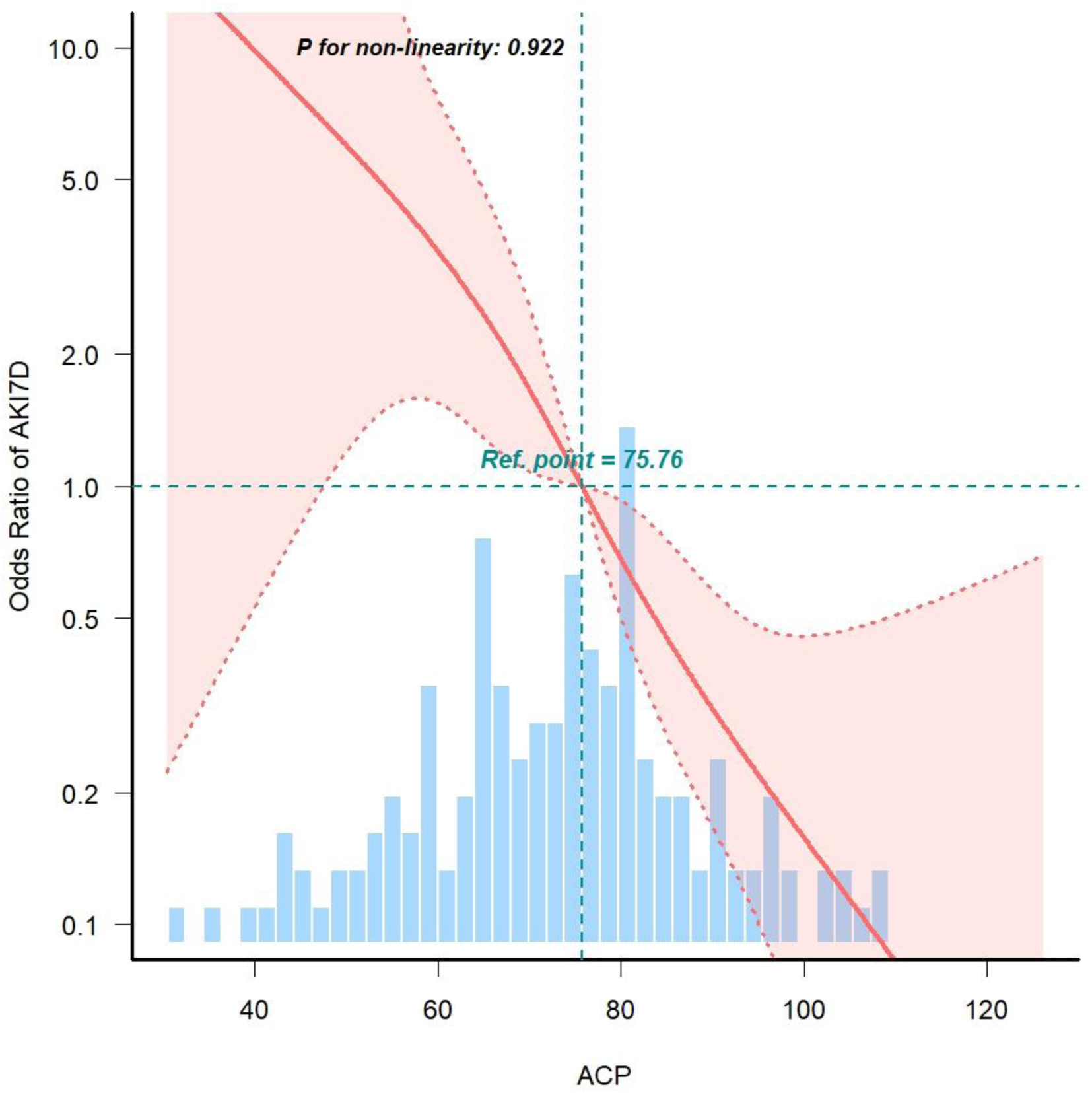
Cubic model of the association between ACP and the risk of SA-AKI.

To further investigate the independent association between them, Cox regression analysis was performed. Results showed that independent risk factors for SA-AKI within 7 days included history of hypertension, SOFA scores, types of pathogens, serum potassium and ACP at enrollment (Table S3-7). Considering the effects of age on renal function, we further included this variable in the multivariate Cox regression analysis. After adjusting for all covariates mentioned above, as a continuous variable, the risk of SA-AKI decreased by 2.1% for every 1% increase in ACP. In addition, as a categorized variable, patients with mild (80% > ACP ≥ 60%) and moderate-to-severe (ACP < 60%) cardiac impairment had a 1.822-fold and 2.364-fold increased risk of SA-AKI within 7 days compared with patients with normal cardiac function, respectively (Table 3).

**Table 3.** Cox proportional hazards regression analyses of SA-AKI according to ACP, CI, CPI and RPP.

| Variables | Crude model |  | Adjusted model 1 |  | Adjusted model 2 |  |
| --- | --- | --- | --- | --- | --- | --- |
|  | HR (95% CI) | *P* value | HR (95% CI) | *P* value | HR (95% CI) | *P* value |
| **Continuous variables** |  |  |  |  |  |  |
| ACP (%) | 0.974 (0.963 - 0.985) | < 0.001 | 0.979 (0.967 – 0.991) | 0.001 | 0.980 (0.968 – 0.991) | 0.001 |
| CI (L/min/m<sup>2</sup>) | 0.774 (0.653 - 0.918) | 0.003 | 0.828 (0.693 – 0.990) | 0.038 | 0.815 (0.682 – 0.974) | 0.024 |
| CPI (W/m<sup>2</sup>) | 0.128 (0.048 - 0.337) | < 0.001 | 0.207 (0.073 – 0.580) | 0.003 | 0.190 (0.069 – 0.526) | 0.001 |
| RPP (mmHg) | 0.973 (0.961 - 0.985) | < 0.001 | 0.978 (0.964 – 0.992) | 0.002 | 0.980 (0.966 – 0.993) | 0.003 |
| **Categorized variables** |  |  |  |  |  |  |
| ACP (%) |  |  |  | 0.014 |  | 0.011 |
| ≥80% | 1.00 (Ref.) |  | 1.00 (Ref.) |  | 1.00 (Ref.) |  |
| 60 - 80% | 2.439 (1.503 - 3.959) | < 0.001 | 1.822 (1.092 – 3.040) | 0.022 | 1.875 (1.134 – 3.100) | 0.014 |
| < 60% | 2.941 (1.665 - 5.194) | < 0.001 | 2.364 (1.300 – 4.296) | 0.005 | 2.368 (1.319 – 4.253) | 0.004 |
| CI (L/min/m<sup>2</sup>) |  |  |  |  |  |  |
| ≥ 3.0 | 1.00 (Ref.) |  | 1.00 (Ref.) |  | 1.00 (Ref.) |  |
| < 3.0 | 1.98 (1.311 – 2.990) | 0.001 | 1.642 (1.073 – 2.512) | 0.022 | 1.721 (1.122 – 2.64) | 0.013 |
| CPI (W/m<sup>2</sup>) |  |  |  |  |  |  |
| ≥0.5 | 1.00 (Ref.) |  | 1.00 (Ref.) |  | 1.00 (Ref.) |  |
| < 0.5 | 1.989 (1.33 – 2.974) | 0.001 | 1.566 (1.026 – 2.390) | 0.038 | 1.623 (1.069 – 2.464) | 0.023 |
| RPP (mmHg) |  |  |  |  |  |  |
| ≥60 | 1.00 (Ref.) |  | 1.00 (Ref.) |  | 1.00 (Ref.) |  |
| < 60 | 2.272 (1.532-3.369) | < 0.001 | 1.966 (1.274 – 3.035) | 0.002 | 1.884 (1.247 – 2.847) | 0.003 |

Adjusted model 1: age, SOFA, serum potassium, history of hypertension, types of pathogens.
Adjusted model 2: age, SOFA, serum potassium, history of hypertension.

Similarly, Cox regression analysis showed that following adjustment for relevant confounders, no matter as a continuous variable or a categorized variable, reduced CI, CPI and RPP emerged as independent risk factors for SA-AKI, too (Table 3).

### Establishment of predictive nomogram model

Based on the multivariate Cox regression model, a nomogram model showing the probability of SA-AKI occurrence within 7 days was established with age, the history of hypertension, SOFA, serum potassium, types of pathogens and ACP at admission as predictors. According to Figure 3A, the cumulative score derived from the individual predictor can be utilized to predict the probability of AKI development in sepsis within a 7-day timeframe. The AUC value of this prediction model was 0.865 (95%CI 0.808 – 0.923, *p* < 0.001) (Figure 3C), and the C-index was 0.800 (95%CI 0.753 - 0.847) (Figure 3B). Given that the majority of sepsis patients do not have pathogen type results at ICU admission, we opted to exclude this variable from the predictive model (Figure 4A). The AUC value of this prediction model was 0.815 (95%CI 0.749 – 0.881, *p* < 0.05) (Figure 4C), and the C-index was 0.780 (95%CI 0.730 - 0.830) (Figure 4B).

**Figure 3.**
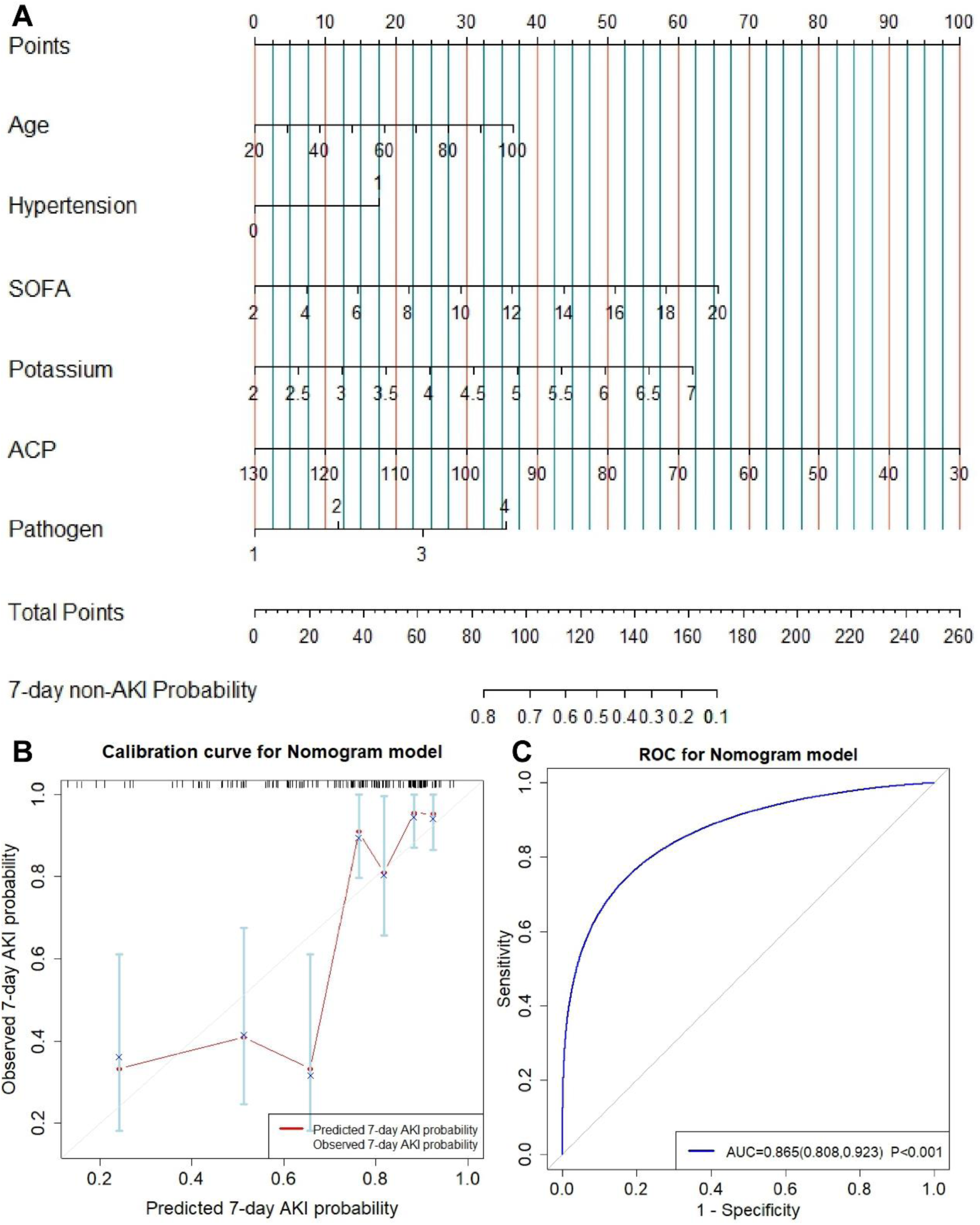
Nomogram model with pathogen types for predicting SA-AKI within 7 days. A. The nomogram model; B. Graphical calibration curve of the nomogram model; C. The ROC curve of nomogram model to predict the incidence of SA-AKI within 7 days.

**Figure 4.**
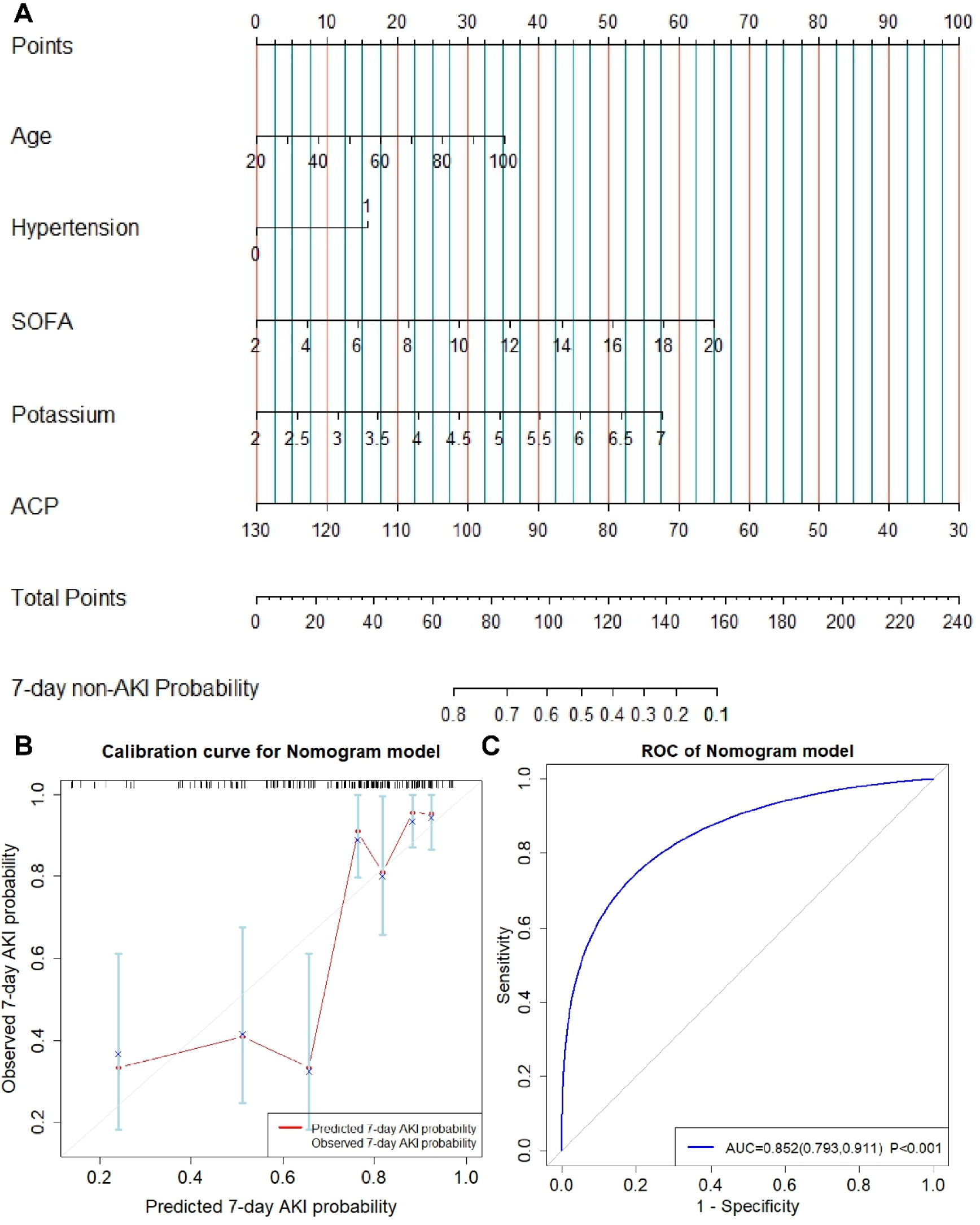
Nomogram model without pathogen types for predicting SA-AKI within 7 days. A. The nomogram model; B. Graphical calibration curve of the nomogram model; C. The ROC curve of nomogram model to predict the incidence of SA-AKI within 7 days.

### Secondary outcome

In the baseline comparison, we found that the hospital mortality increased with the aggravation of myocardial impairment, in addition, the patients with moderate to severe cardiac function impairment had the highest in-hospital mortality (65.5%) and the shortest length of ICU stay (Table S1). Kaplan-Meier curve showed that the patients in moderate-severe impairment had higher hospital mortality than patients in the normal group (Log-rank test: *p* < 0.05, Figure 5).

**Figure 5.**
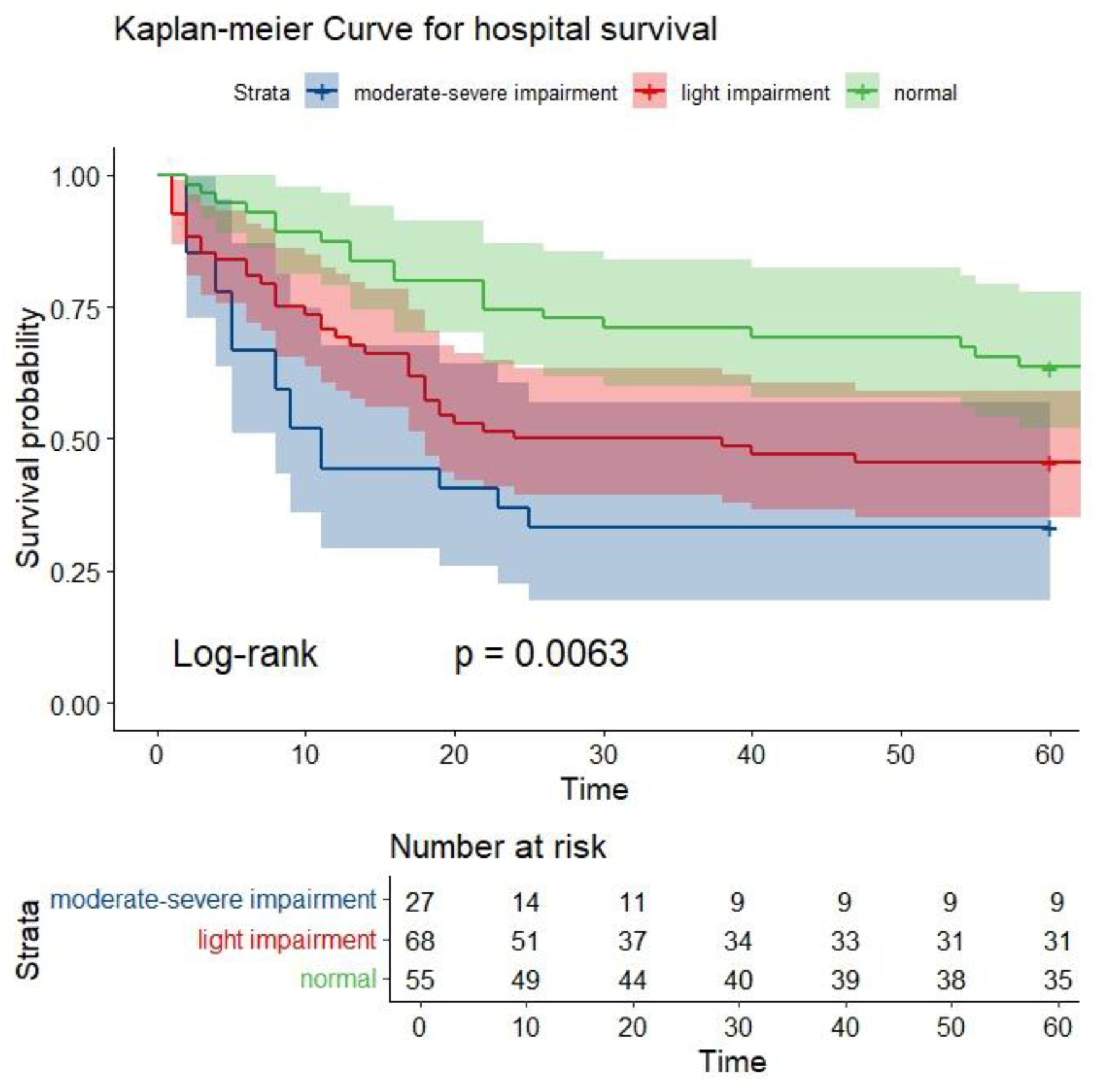
Kaplan-Meier Survival Curves of hospital mortality in patients with different degrees of cardiac function impairment.

## Discussion

In fact, as early as 1951, Ledoux et al. have pointed out that cardiac insufficiency can cause impaired kidney function^[29]^. In 2008, Ronco et al. further emphasized the interaction between heart and kidney, and proposed that cardiorenal syndrome (CRS) is a clinical syndrome in which acute or chronic dysfunction of one of the heart or kidney leads to acute or chronic impairment of the other organ^[30]^. Sepsis is the main and most common cause of acute Type 5 CRS, which has a high mortality. In our study cohort, 67.3% of patients with sepsis developed AKI. Patients with decreased ACP had a significantly higher incidence of SA-AKI within 7 days than those with normal ACP. Our study also suggested that the decrease of ACP is an independent risk factor for SA-AKI within 7 days. Therefore, our findings once again validate the interaction of cardio-renal function in sepsis.

The pathogenesis of Type 5 CRS is relatively complex. It is believed that it is related to renal hypoperfusion due to decreased cardiac output caused by cardiac insufficiency on the one hand, and increased renal venous pressure caused by continuous venous congestion on the other hand^[31]^. In recent years, ACP has been considered as a potential indicator for diagnosing cardiac function impairment and associated with the severity and short-term prognosis of patients with sepsis^[32–36]^. Our previous study has shown that a strong linear correlation between ACP and CI as well as CPI^[37]^. Moreover, the relevant measures used to calculate ACP are obtained from Pulse indicator continuous cardiac output (PiCCO), which have clinical utility. Therefore, the application of ACP in forecasting the occurrence of AKI had a theoretical basis.

In fact, previous studies have demonstrated the relationship between traditional indicator CI and AKI. Luo et al. showed that an improved CI was a protective factor for renal function in patients with septic shock^[38]^. However, several studies have found that patients with septic shock often exhibit pseudonormalization or even elevation of CI due to a dramatic decrease in SVR^[39]^. Therefore, only focusing on CI may lead to an underestimation of the risk of AKI occurrence. Our retrospective study also verified that CI had a low sensitivity in predicting the occurrence of SA-AKI within 7 days. The predictive value of ACP for SA-AKI within 7 days was significantly better than CI, with better specificity and sensitivity, and the cut-off value was also very close to the threshold of 80% defined by cardiac function decline. Therefore, we believe that early monitoring of ACP is conducive to predicting the risk of SA-AKI.

CPI is considered to be an indicator that helps to identify the residual systolic reserve function in patients with heart failure^[40]^. It makes up for the deficiency of CI to a certain extent by modifying MAP, but there is limited studies on the relationship between CPI and AKI. Our study found that low CPI has a high sensitivity in predicting the occurrence of SA-AKI. RPP, which reflects renal blood perfusion, was less sensitive than ACP in predicting SA-AKI, but had a higher specificity (87.8%). RPP represents the transrenal pressure gradient and is the difference between inflow pressure (MAP) and out flow pressure [CVP or intra-abdominal pressure (IAP)]^[28]^. The predictive cut-off value of RPP was 60 mmHg, which led us to consider that for sepsis patients with a CVP > 5mmHg, maintaining a target MAP of 65mmHg may not be sufficient in preventing AKI occurrence.

From the clinical perspective, the nomogram model in this study has strong rationality and clinical practicability. The factors in this model are clinical data easy to obtain, and the application is relatively simple. Numerous studies have reported the effects of age and high blood pressure on kidney function. SOFA is an indicator of disease severity and has often been used in previous studies to predict AKI^[41]^. Blood potassium level may be a side reflection of renal excretion function. As for the relationship between fungal infection and AKI, there is no clear report at present, and further research is needed to confirm it. Statistically, the AUC of the nomogram model with pathogenic species was up to 0.865, indicating that the model had good differentiation. In addition, the C-index of the model reaches 0.800, which also indicates that the model has good calibration degree. Therefore, the SA-AKI prediction model established in this study not only has clinical practicability, but also has high accuracy.

Furthermore, our study demonstrated that patients with myocardial impairment (regardless of mild to severe) had a higher risk of in-hospital death than normal patients. As far as we know, ACP is associated with the prognosis of patients with sepsis^[32, 34]^, and the results of this study are consistent with these findings.

It’s is important to note that ACP has the limitation of not accounting for preload. Therefore, preload independence must be assessed correctly before standardizing ACP measurement. In our study, there was no significant difference in GEDI between the groups, suggesting that all patients had received early and adequate fluid resuscitation to minimize the impact of preload.

There were several limitations must be considered in our study. First, this was a retrospective observational cohort study. Some data were missing, such as troponin and brain natriuretic peptide, which could not be included in the analysis. However, we excluded patients with an acute myocardial infarction during the same hospitalization at the time of screening, and the small percentage of patients with chronic heart failure in our study, which minimized the bias in the results. Second, our finding was based on data obtained from patients undergoing PiCCO monitoring, which might lead to selection bias. Third, the sample size was small, and the confidence intervals of both tails of the fitting curve were large, and large prospective multicenter studies are needed to further verify the stability of the results. Finally, our study followed patients for a short time and did not pay much attention to the long-term prognosis.

### Conclusions

There was a negative correlation between ACP and the occurrence of SA-AKI. In addition, ACP, which may be superior to traditional cardiac function indicators, had the potential value of early prediction of SA-AKI. Decreased ACP might increase the risk of in-hospital mortality in patients with sepsis.

## Data Availability

The dataset file has been uploaded.

## Supplementary materials

The supplemental files contain the tables needed to be displayed in the result section.

## Acknowledgements

None.

